# Membranous Nephropathy: Muti-omics Mendelian randomization and colocalization analysis

**DOI:** 10.1101/2023.11.21.23298831

**Authors:** Zhihang Su, Wen Liu, Zheng Yin, Di Wu, Yuan Cheng, Qijun Wan

## Abstract

**Background:** The currently available medications for treating membranous nephropathy (MN) still have unsatisfactory efficacy in inhibiting disease recurrence, slowing down its progression, and even halting the development of end-stage renal disease. There is still a need to develop novel drugs targeting MN.

**Methods:** We utilized summary statistics of MN from the Kiryluk Lab and obtained plasma proteins from Zheng et al., Decode, and UKBioBank and gene data from eQTLgen and GTEX. We performed a two-sample Bidirectional mediation Mendelian randomization analysis, SMR analysis, HEIDI test, Bayesian colocalization, phenotype scanning, external validation, mediation analysis, drug bank analysis, and protein-protein interaction network.

**Results:** The Mendelian randomization analysis uncovered 8 distinct proteins associated with MN after False Discovery Rate multiple correction. Proteins associated with an increased risk of MN in plasma include ABO [(Histo-Blood Group Abo System Transferase) (WR OR = 1.116, 95%CI:1.047-1.190, FDR=0.090, PPH4 = 0.795)], VWF [(Von Willebrand Factor) (WR OR = 1.412, 95%CI:1.157-1.725, FDR=0.018, PPH4 = 0.816)] and CD209 [(Cd209 Antigen) (WR OR = 1.187, 95%CI:1.074-1.312, FDR=0.090, PPH4 = 0.795)], and proteins that have a protective effect on MN: HRG [(Histidine-Rich Glycoprotein) (WR OR = 1.814, 95%CI:1.345-2.445, FDR=0.018, PPH4 = 0.797)], CD27 [(Cd27 Antigen) (WR OR = 0.785, 95%CI:0.681-0.904, FDR=0.018, PPH4 = 0.797)], LRPPRC [(Leucine-Rich Ppr Motif-Containing Protein, Mitochondrial) (WR OR = 0.790, 95%CI:0.688-0.907, FDR=0.090, PPH4 = 0.797)], TIMP4 [(Cd27 Antigen) (WR OR = 0.666, 95%CI:0.527-0.840, FDR=0.090PPH4 = 0.833)] and MAP2K4 [(Metalloproteinase Inhibitor 4) (WR OR = 0.815, 95%CI:0.723-0.919, FDR=0.090, PPH4 = 0.797)]. None of these proteins exhibited a reverse causal relationship. Bayesian colocalization analysis provided evidence that all of them share variants with MN. In external validation, ABO, CD27, HRG, MAP2K4, TIMP4, and VWF showed significant mediation results. We identified type 1 diabetes, trunk fat, and asthma as having intermediate effects in these pathways. We discovered several genes that are causally related to MN.

**Conclusions:** Our comprehensive analysis indicates a causal effect of ABO, CD209, CD27, HRG, LRPPRC, MAP2K4, and TIMP4 at the genetically determined circulating levels on the risk of MN. These proteins have the potential to be a promising therapeutic target for the treatment of MN. We identified type 1 diabetes, trunk fat, and asthma as having intermediate effects in these pathways. We discovered several genes that are causally related to MN.

## 1 Introduction

Membranous Nephropathy (MN) is a common chronic autoimmune disease.

MN may occur in both sexes and all ethnic groups. It is widely acknowledged that MN is the most common cause of adult nephrotic syndrome(1). The prognosis of MN is challenging to predict, as some patients may experience spontaneous remission while others progress to end-stage renal disease(2). Current mainstream treatments for MN include cyclophosphamide, glucocorticoids, rituximab, and calcineurin inhibitors(3). In contrast to newly developed therapies targeting BAFF, plasma cells, and complement(4), human circulating proteins play a crucial role in various biological processes and serve as primary drug targets(5). The relevant literature has shown that protein drug targets with genetic associations supporting their connection to diseases are more likely to become drug targets and gain market recognition compared to targets discovered by chance(6,7). In recent times, the application of Mendelian randomization (MR) analysis has gained significant traction in identifying potential therapeutic targets and repurposing drugs(8). As an approach within genetic instrumental variable analysis, MR employs single nucleotide polymorphisms (SNPs) as instrumental variables (IVs) to estimate the causal relationship between exposures and outcomes(9). Compared to traditional observational studies, MR has the ability to mitigate the impacts of type I errors and confounding factors(10). As a result of advancements in large-scale genomic and proteomic technologies, the utilization of MR-based strategies has enabled the discovery of promising targets for therapeutic interventions in a variety of disorders, including Heart Failure(11), multiple sclerosis(12), and inflammatory bowel disease(13). However, no MR study reports have integrated GWAS of MN and protein quantitative trait loci (pQTL) data to date. In the present investigation, our focus was to ascertain plasma as a viable candidate for therapeutic intervention in the case of MN. Figure 1 depicts the study design. Initially, we utilized MN data from Kiryluk Lab and plasma pQTL data to discern plausible causal plasma proteins associated with MN.(14) Next, we further validated the initial findings using Steiger filtering, Bayesian colocalization analysis, and phenoscanner. In a subsequent step, we constructed an interaction network linking potential protein targets with MN drug targets. Lastly, for reinforcement of our conclusions, we also conducted external validation. We assessed the mediating factors between these significant proteins and MN. Finally, we analyzed the relationship between the genes of these significant proteins and MN using expression quantitative trait Loci (eQTL) data, GTEX, the smr method, and the HEIDI test.

**Figure 1:**
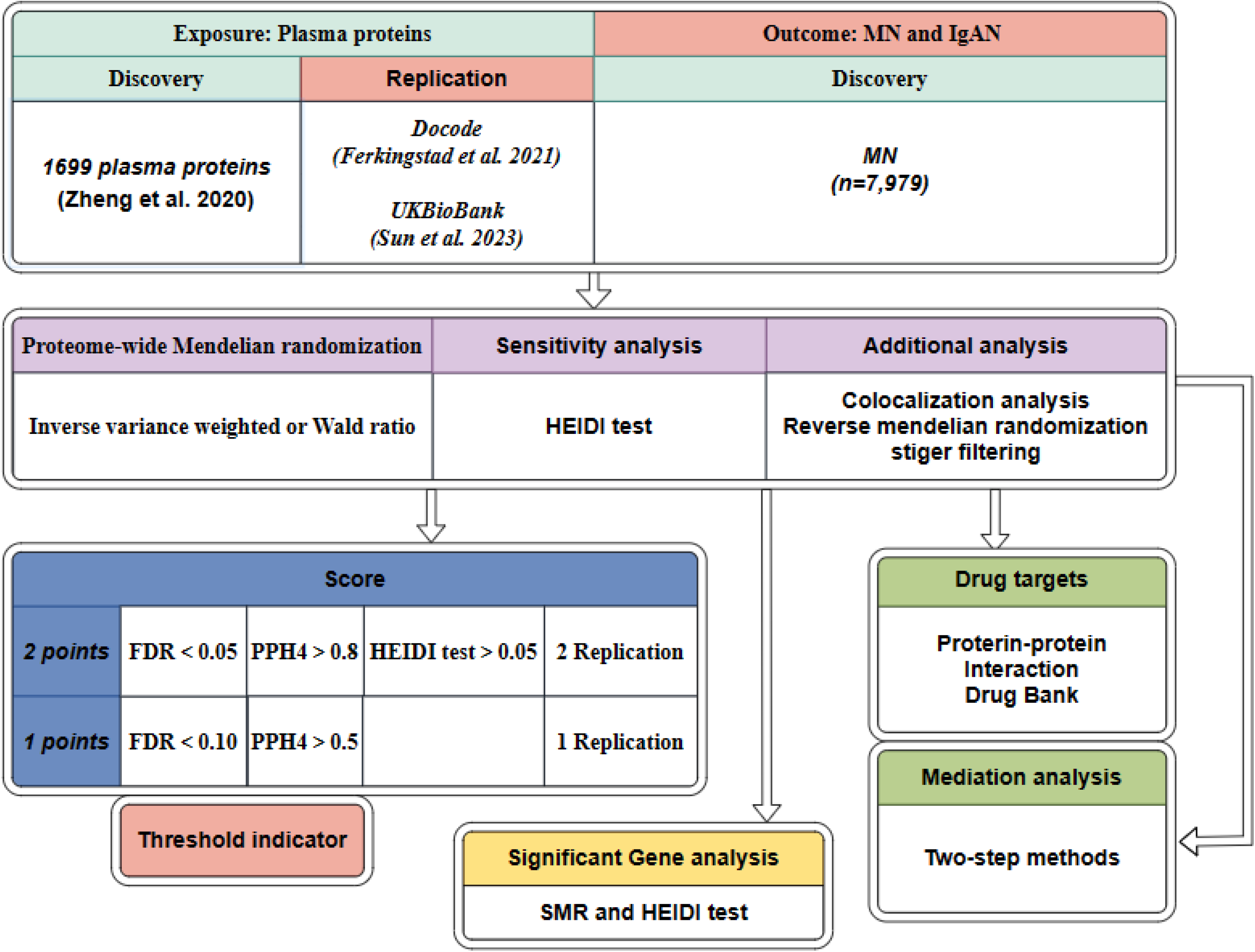
Study design

## 2 Methods

### 2.1 Plasma protein quantitative trait loci

We incorporated pQTLs that fulfilled the subsequent set of criteria: ① demonstrating a significant correlation at the genome-wide level (P < 5e-8); ② being situated outside the major histocompatibility complex (MHC) region (chr6, 26-34Mb); ③ demonstrate distinct associations [linkage disequilibrium (LD) clustering r^2 < 0.001]. In the preliminary analysis, we extracted plasma pQTL data from public research,(14) which consolidated findings from five public studies(15–19). With regard to the aforementioned selection criteria, a total of 2,113 cis and trans SNPs associated with 1,699 proteins were included. The aforementioned databases have been concurrently utilized in previous studies, alleviating concerns regarding their reliability.(12) Each database will be treated as an individual sample, and we will conduct MR on each of them. Plasma pQTL data of external validation was retrieved from recent study. Decode encompassed 35,559 participants enabling the quantification of a total of 4,907 plasma proteins(20). UKB-PPP encompassed 54,219 participants, enabling the quantification of a total of 2,923 plasma proteins(21). The above databases are all derived from European populations.

### 2.2 Genome-wide association study data of Membranous nephropathy

Aggregate statistical data of MN, including individuals of European ancestry (Sample size of Cases=2150, Sample size of Control=5829, size of SNPs=5,327,688), were retrieved from the Kiryluk Lab.(22) All cases included in the final database were diagnosed as idiopathic membranous nephropathy using the gold standard method of renal biopsy. Furthermore, potential causes of secondary membranous nephropathy, such as medication, malignancies, infections, and autoimmune diseases, were systematically ruled out. Nonetheless, the study did not present a detailed description of the target antigens in the patients.

### 2.3 Mendelian randomization analysis

#### 2.3.1 Selection of Instrumental Variables

The IVs must meet the mentioned assumptions: relevance, exogeneity, and independence.

i. The IVs must exhibit a strong association with plasma protein. First, we selected SNPs falling beneath the locus-wide significance cutoff (5 × 10^−8^).
ii. The clumping processes were executed to maintain the IVs’ independence and remove the linkage disequilibrium (LD) effect. Linkage disequilibrium (r^2^) was set at 0.001, and genetic distance was set at 10,000 kb.
iii. F = R2(n - k - 1)/ k (1 - R2), where R2 is the exposure variance calculated based on the data within the dataset. The number of IVs is k, and the samples are n. When F-statistic<10, it indicates weak IVs, and they will be excluded.

Within this study, proteins were investigated as the primary variable of interest, representing the exposure, while MN served as the focal point of analysis, representing the outcome. The Wald ratio (WR) was utilized when it came to one SNP(23). When two or more SNPs were accessible (the numbers of SNPs≥2), we utilized the inverse variance weighted (IVW) method, followed by assessments of heterogeneity and horizontal pleiotropy(24). In the process of conducting Mendelian randomization (MR), in cases where effect allele frequency data is missing, we employ data from the 1000 Genomes project (HG19/GRCh37) as a substitute. The amplified risk of MN was indicated as odds ratios (OR) per one-unit change in plasma protein levels, corresponding to a standard deviation (SD) increase.

### 2.4 Reverse causality detection and sensitivity analysis

In the presence of a sufficient number of SNPs, we will conduct a bidirectional MR analysis. If an adequate number of SNPs is not available, we will resort to Stiger filtering as an alternative, which is also a directional testing method(25). Unlike most methods based on GWAS data, the Heterogeneity in Dependent Instruments (HEIDI) test is used to distinguish between pleiotropy and linkage models. Associations with a HEIDI test p-value < 0.05 are considered to be caused by pleiotropy and are therefore removed from further analysis. The SMR software tool (SMR v1.0.3) was used for SMR analysis.

### 2.5 Bayesian colocalization analysis

Colocalization analysis was employed to evaluate the likelihood of two traits sharing a common causal variant. As previously mentioned, the posterior probabilities of five hypotheses were utilized to provide detailed supplementation to the results of colocalization analysis. Specifically, the first hypothesis (PPH0) indicates “No association with either trait.” The second hypothesis, posterior probabilities of hypotheses (PPH1), represents, in this gene segment, there is only an association with trait 1 but not with trait 2. Lastly, the third hypothesis (PPH2) signifies “Association with trait 2, but not with trait 1.” In our research, we assessed the posterior probabilities of hypotheses 3 and 4 (PPH3, PPH4).(26) In hypothesis 3, exposure (plasma protein) and outcome (MN) are linked to the genomic region through different SNPs, while in hypothesis 4, the protein and MN are both linked to the genomic region through a shared SNP. By employing the coloc.abf algorithm, we defined genes as having gene-based colocalization evidence when PPH4 > 80%.

### 2.6 Phenoscanner

We also employed Phenoscanner to uncover the connections of identified pQTLs with other traits. SNPs were classified as having horizontal pleiotropy when they exhibited connections with established risk factors for MN, encompassing metabolic traits, proteins, or clinical characteristics.

### 2.7 Mediation analysis

We conducted a mediation analysis on significant proteins using a two-step Mendelian randomization approach to identify the mediating factors. The overall influence of exposure on the outcomes was categorized into direct effects and indirect effect(27). Specifically, the total effects of the identified plasma proteins on MN in this study encompassed two components: 1) the direct effects of the identified plasma proteins on MN, calculated through primary MR analysis; and 2) using the product method to estimate the indirect effects mediated by the identified risk factors. Standard errors (SE) and confidence intervals (CI) were determined employing the delta method.

### 2.8 SMR analysis

We further performed SMR analysis to identify genes that may have a causal relationship with MN. The data used for analysis were sourced from eqtlgen and GTEX (28). The HEIDI test was concurrently utilized to distinguish between pleiotropy and linkage models.

### 2.9 Protein-protein interaction network

In the plasma analysis, we investigated proteins potentially associated with MN. We primarily employed MR analysis with a significance level of FDR < 0.10 and directional tests. Furthermore, we explored the target proteins of drugs for MN available in the market. We identified these targeted drugs through a comprehensive review of the literature and by searching the DrugBank database (<u>DrugBank | Powering health insights with structured drug data</u>).(29,30) Additionally, we conducted a search for drugs targeting the potential pathogenic proteins we identified. Subsequently, protein-protein interaction (PPI) networks were employed to investigate the interplay among the proteins and the relevant targets. All PPI analyses were executed utilizing version 12.0 of the STRING database [<u>STRING: functional protein association networks (string-db.org)</u>],(31) with a predefined minimum threshold set at 0.5.

## 3 Results

### 3.1 Mendelian randomization analysis

Following the adherence to the principles of instrumental variable selection, a comprehensive set of 1057 IVs were included in the analysis. After False Discovery Rate multiple correction, employing a nominal significance threshold (P<0.10), the MR analysis s identified 8 protein-MN associations. We discovered plasma proteins associated with an increased risk of MN: ABO [(Histo-Blood Group Abo System Transferase) (WR OR = 1.12, 95%CI:1.05-1.19, FDR=0.09, PPH4 = 0.80)], VWF [(Von Willebrand Factor) (WR OR = 1.412, 95%CI:1.16-1.73, FDR=0.02, PPH4 = 0.82)] and CD209 [(Cd209 Antigen) (WR OR = 1.19, 95%CI:1.07-1.31, FDR=0.09, PPH4 = 0.80)] and proteins that have a protective effect on MN: HRG [(Histidine-Rich Glycoprotein) (WR OR = 1.81, 95%CI:1.35-2.45, FDR=0.02, PPH4 = 0.80)], CD27 [(Cd27 Antigen) (WR OR = 0.79, 95%CI:0.68-0.90, FDR=0.02, PPH4 = 0.80)], LRPPRC [(Leucine-Rich Ppr Motif-Containing Protein, Mitochondrial) (WR OR = 0.79, 95%CI:0.69-0.91, FDR=0.09, PPH4 = 0.80)], TIMP4 [(Cd27 Antigen) (WR OR = 0.67, 95%CI:0.53-0.84, FDR=0.09, PPH4 = 0.83)] and MAP2K4 [(Metalloproteinase Inhibitor 4) (WR OR = 0.82, 95%CI:0.72-0.92, FDR=0.09,PPH4 = 0.80)]. (Table 1). Additionally, replication of these proteins can be obtained in the decode and UK Biobank datasets (Table1).

### 3.2 Sensitivity analysis and Reverse causality detection for MN causal proteins

During the Phenoscanner process, no confounding factors were found. The preliminary analysis did not detect any heterogeneity or horizontal pleiotropy in the analyzed proteins. In the reverse causal analysis (with MN as the exposure and plasma proteins as the outcome), the aforementioned positive proteins did not exhibit a causal relationship with membranous nephropathy. In addition, we resorted to directional testing to evaluate causality. Furthermore, Steiger filtering provided additional assurance regarding the presence of directional abnormalities. Steiger filtering consistently showed the correct directionality, and the significance level was P<0.05. In the HEIDI test, ABO, HRG, and TIMP4 passed the test, while CD209 did not pass the test(Table 2). The remaining four proteins have missing data.

### 3.3 Bayesian co-localization analysis

Through colocalization analysis,All significant proteins exhibit co-localization with moderate or higher strength[strong Bayesian colocalization (PPH4>0.8) and Moderate-strength Bayesian colocalization (0.5 < PPH4 < 0.8)]. This indicates that these significant plasma proteins share the same variants with MN(Table 1).

### 3.4 SMR analysis for gene

After excluding the effects of heterogeneity and horizontal pleiotropy through the HEIDI test, we identified a significant causal relationship between numerous genes and membranous nephropathy based on gene prediction. Among them, VWF (ENSG00000110799) in the pqtl was validated through SMR analysis using the eQTLgen database (Supplementary Table S1 and S2).

### 3.5 The correlation among causal proteins and current drug targets for MN

The PPI network unveiled an interrelationship involving three proteins (CD27) and an existing drug target for MN (CD20 targeted by rituximab). Using STRING, the identified reliable protein interaction axes include MS4A1-CD27-NCAM1, and MS4A1-NCAM1-CD27 (where NCAM1 has been previously confirmed as a pathogenic antigen for MN) (Supplement Table S3). Therefore, we should pay more attention to the relationship between MS4A1 and CD27. For unknown proteins, CD27 is linked to the B lymphocyte antigen CD20 (MS4A1), which serves as the focus for rituximab. Furthermore, STRING analysis unveiled physical interactions between the aforementioned proteins and MS4A1, suggesting their close proximity without necessarily being in direct contact. The phenotype scan revealed that the aforementioned correlated proteins remained unaffected by other phenotypes, as all the phenotypes examined were unrelated to MN (Supplement Table S4).

### 3.6 Mediation analysis

Our two-step analysis unveiled multiple factors that function as mediators in the causal pathway. These include adult-onset asthma, beef intake, diabetes or endocrine disease, E-selectin levels, trunk fat, type 1 diabetes with other specified/multiple/unspecified complications, and type 1 diabetes, all of which have the potential to act as mediators in the associations between proteins and diseases(Supplementary table S5).

## 4 Discussion

To our knowledge, this study represents the first attempt to integrate plasma proteomics data with MR and Bayesian colocalization approaches using many databases to investigate the potential involvement of specific proteins in MN. Furthermore, the relationship between genes and MN was analyzed using SMR analysis. Based on genetic prediction, ABO, CD209, CD27, HRG, LRPPRC, VWF, MAP2K4, and TIMP4 were found to have a causal relationship with MN. Through a series of analyses, we ultimately identified ABO, HRG, and TIMP4 as high-priority potential drug targets for MN. Although CD27, CD209, VWF, LRPPRC, and MAP2K4 had a lower priority, they still hold suggestive significance. A one standard deviation (SD) change in the concentration of plasma proteins in the blood is associated with an odds ratio (OR) effect on MN. Our research in proteomics holds significance, as proteins, acting as effectors in numerous biological processes,(32) are widely regarded as drug targets, unlike previous studies that primarily focus on single phenotypes or even precise dual-disease MR, which are typically gene-centric analyses susceptible to the influence of gene-gene and gene-environment interactions. Taking advantage of the rapid advancement in high-throughput sequencing, we integrated genome-wide association studies (GWAS) with proteomics, enabling the integration of genetic and protein-level information. By utilizing MR and employing a series of validation methods, we have newly identified several promising proteins, thereby augmenting their potential as targets for therapeutic interventions. Hence, in order to identify innovative targets for drug intervention in MN, we utilized a comprehensive analysis to evaluate the causal involvement of proteins in MN. Causal relationships identified through MR may include reverse causality, horizontal pleiotropy, or genetic confounding induced by linkage disequilibrium (LD). Therefore, in this study, we adhered to the three fundamental principles of instrumental variable selection in Mendelian randomization analysis. We utilized IVs that exhibited strong correlations with plasma proteins. We excluded IVs that demonstrated linkage disequilibrium during the implementation of MR. The reverse Mendelian randomization analysis revealed that there is no reverse causal relationship between these proteins and MN. We utilized cis and trans pQTLs as IVs as they are implicated in the regulation of gene expression at both the transcriptional and translational levels. Cis-pQTLs refer to variants located proximal to the gene encoding the protein under study, while trans-pQTLs represent distal regulatory elements that influence proteins through often unknown mechanisms.(33,34) Trans-pQTLs offer valuable insights into molecular connections in human biology. Additionally, Bayesian colocalization was employed to eliminate bias introduced by LD.(35) Using a posterior probability threshold of 0.5, it was considered as moderate to high co-localization strength, and we identified 8 proteins that potentially share the same variants associated with MN. Nevertheless, these associations alone do not provide a complete explanation for the connection between the identified proteins and MN. Fortunately, Phenoscanner analysis unveiled associations of the IVs for these proteins with various phenotypes such as Duodenal ulcer, APTT, and thrombosis. These diverse phenotype associations are unrelated to MN, making it unlikely that the results are biased. Therefore, ABO, CD209, CD27, HRG, LRPPRC, VWF, MAP2K4, and TIMP4 may serve as potential drug targets for MN, with particular emphasis on ABO, HRG and TIMP4. We also conducted network and database searches for targeted drugs against CD27, such as MK-5890 (monoclonal antibody),(36) Varlilumab (monoclonal antibody),(37), and CDX-527 (bispecific antibody).(38) These drugs hold promise as novel targeted therapies for the treatment of MN.

ABO, Histo-blood group ABO system transferase is a critical protein involved in determining the ABO blood group system, which encompasses three carbohydrate antigens: A, B, and H. This protein facilitates the transfer of specific sugar molecules onto the surfaces of red blood cells, thereby determining an individual’s blood type within the ABO system. This may suggest that blood type has an impact on the risk of developing MN.

CD209,CD209 antigen,The pathogen-recognition receptor is expressed on the surface of immature dendritic cells (DCs) and plays a crucial role in initiating the primary immune response. It is believed to mediate the endocytosis of pathogens, which are then degraded in lysosomal compartments. Subsequently, the receptor returns to the cell membrane surface, presenting pathogen-derived antigens to resting T-cells through MHC class II proteins, thereby initiating the adaptive immune response.

CD27 functions as a receptor for CD70/CD27L and may play a crucial role in the survival of activated T-cells. The MR result suggests that for every one standard deviation increase in circulating levels of CD27, there is a 0.784% decrease in the risk of MN. It is implicated in cellular apoptosis through its interaction with SIVA1.(39) CD27 exhibits protein-protein interactions with CD20 and NCAM1. While CD20-targeted therapy, rituximab, has become the mainstream treatment for MN. The combination of PPI network analysis and MR revealed that therapeutic agents targeting CD27 have exhibited promising progress and evaluation in clinical trials, in contrast to the other identified targets in our study. Currently, targeted drugs against CD27, such as MK-5890 (monoclonal antibody), Varlilumab (monoclonal antibody), and CDX-527 (bispecific antibody), hold potential as new targeted therapies for the treatment of MN. Based on our research findings, we hypothesize that these antibodies may have an impact on MN, and therefore, we recommend future clinical studies to further investigate this hypothesis.

HRG, also known as Histidine-Rich Glycoprotein. Plasma glycoprotein can bind to various ligands, such as hemoglobin, acetylated hepari and plasminogen.

LRPPRC,The leucine-rich PPR motif-containing protein is involved in RNA metabolism in both the nucleus and mitochondria. In the nucleus, it binds to poly(A) mRNAs and is associated with mRNA maturation and export. In mitochondria, it binds to poly(A) mRNA and affects the translation and stability of cytochrome c oxidase subunits.

VWF, also known as Von Willebrand Factor. It plays a vital role in hemostasis and serves as a carrier for clotting factors. Upon blood vessel damage, a significant number of platelets adhere to collagen fibers, forming a clot, where VWF acts as a mediator in hemostasis. The binding of VWF factor to clotting factor VIII stabilizes and regulates its activity.

Dual specificity mitogen-activated protein kinase kinase 4 (MAP2K4) is a protein kinase that plays a crucial role in the MAP kinase signal transduction pathway, specifically in the stress-activated protein kinase/c-Jun N-terminal kinase (SAP/JNK) signaling pathway. It directly activates MAPK8/JNK1, MAPK9/JNK2, and MAPK10/JNK3, making it one of the few known kinases to do so.

Metalloproteinase inhibitor 4 (TIMP-4) forms complexes with metalloproteinases, specifically collagenases, and inactivates their activity by binding to the catalytic zinc cofactor.

In conclusion, we utilized proteomics and MR analysis to identify drug targets for MN. Unlike previous studies that focused solely on the genetic level, our analysis incorporated both genetic and protein levels. Considering the limited availability of targeted drugs for the treatment of MN, we have successfully identified several potential therapeutic targets that require further investigation in the future.

There are several limitations in our research. Firstly, the plasma protein data we utilized was obtained from a combination of multiple proteomic studies, which may employ varying standards for protein measurement. Nevertheless, we selected data that matched circulating protein data with GWAS. Secondly, most proteins are associated with only one SNP, indicating either a lack of trans-pQTL or cis-pQTL. Secondly, the data we employed in our experiments exclusively represented European populations, necessitating further research to generalize the findings to other ethnicities. Thirdly, we did not observe any other proteins in the PPI network that are associated with CD20, except for CD27. This suggests that these proteins may be independent of CD20 and represent alternative potential targets. Additional research is warranted in this regard.

## 5 Data availability

Plasma pQTLs data was derived via public research. (14,20,21) MN data was retrieved from Kiryluk Lab.(22) We used R language version 4.3.0 for our analysis. In R language, we utilized the “coloc (https://github.com/chr1sallace/coloc.git)” and “TwoSampleMR (https://github.com/MRCIEU/TwoSampleMR.git)”, “MendelR” and “locuscompare” packages for our analysis.(40)

## 7 Author Contributions

The main efforts were jointly accomplished by Zhihang Su and Qijun Wan. Wen Liu, Zheng Yin, Di Wu, and Yuan Cheng participated in revising the manuscript.

## 9 Acknowledgments

Sincere thanks to all participants who participated in the production of the public database.

